# Biomarker-Informed Interpretation of Dyadic Cognitive Function Index Scores in Cognitively Unimpaired Older Adults

**DOI:** 10.64898/2026.07.27.26359000

**Authors:** Ambre Mounié, Kenichiro Sato, Saki Nakashima, Yoshiki Niimi, Takeshi Iwatsubo

**Affiliations:** Dementia Inclusion and Therapeutics, The University of Tokyo Hospital, Tokyo, Japan; Cognitive Sciences, Université Côte d’Azur, Nice, France; Unit for Early and Exploratory Clinical Development, The University of Tokyo Hospital, Tokyo, Japan; Department of Neurology, The University of Tokyo Hospital, Tokyo, Japan; National Center of Neurology and Psychiatry, Tokyo, Japan

**Keywords:** Alzheimer’s disease, Cognitive Function Index, amyloid PET, tau PET, plasma p-tau217

## Abstract

**Introduction:** Participant- and study partner-reported Cognitive Function Index scores may provide complementary information, but it remains unclear whether Alzheimer’s disease biomarkers are associated with CFI scores across reporters, reporter-specific imbalance, or both.

**Methods:** Using A4/LEARN screening data (screening sample, N = 1,686; primary dyadic analytic sample, N = 1,682; CDR global score = 0), we jointly modeled participant-reported (CFI-PT) and study partner-reported (CFI-SP) scores in long format to evaluate biomarker associations with CFI scores and biomarker × reporter interactions. As a secondary analysis, we compared tau PET with plasma p-tau217.

**Results:** In an A4/LEARN-adapted regional extent model, reporter balance varied across amyloid regional extent categories, with the largest participant-leading contrast observed in the exploratory restricted early cortical subgroup. Tau PET was associated with higher CFI scores, but this association did not differ detectably between reporters. Plasma p-tau217 showed no clear CFI association or reporter-specific interaction in the A4-derived, amyloid-enriched subset.

**Discussion:** Amyloid regional extent and tau PET were associated with different features of dyadic CFI data: reporter balance and CFI burden across reporters, respectively. Joint interpretation of CFI-PT and CFI-SP may support biomarker-informed interpretation of the instrument, although the cross-sectional findings require replication and longitudinal validation.

## Introduction

Alzheimer’s disease (AD) is characterized by a long preclinical phase during which pathological changes accumulate before cognitive symptoms emerge [Caselli2013; Sperling2011]. Among the earliest clinical manifestations of this underlying pathology are alterations in self-awareness of memory functioning [Vannini2017a]. Anosognosia, defined as a lack of awareness of an individual’s own cognitive deficits [Vannini2017a], is a common feature of AD [Reed1993].

Conversely, hypernosognosia is the heightened awareness of subtle memory changes in the absence of objective cognitive deficits [Vannini2017a; Cacciamani2021]. These observations have led to the hypothesis that awareness-related complaint patterns may vary across the preclinical and prodromal stages of AD [Vannini2017b]. Prior work suggests that cognitively normal (CN) individuals with amyloid pathology (Aβ+) demonstrated heightened memory self-awareness (hypernosognosia), while mild cognitive impairment (MCI) Aβ+ patients tended to overestimate their memory functioning (anosognosia) [Vannini2017a]. By contrast, MCI individuals without amyloid pathology demonstrated close to full insight into their memory functions [Therriault2018]. Alterations in memory self-awareness may also help discriminate individuals likely to progress to AD dementia [Hanseeuw2020]. These awareness-related observations provide an important context for interpreting participant and study partner reports, although disagreement between reporters is not itself a direct measure of cognitive awareness.

A common approach to characterizing differences in perceived cognitive change is to compare participant-reported (PR) and study partner-reported (SPR) cognitive concerns. In the present study, this dyadic framework was operationalized using the Cognitive Function Index (CFI) [Walsh2006; Amariglio2015]. The relative importance of participant- and study partner-reported concerns may shift along the AD continuum, with self-report potentially reflecting awareness alterations early on and study partner report becoming more sensitive to pathology at later stages [Buckley2015; Amariglio2020]. Specifically, higher amyloid and tau burden at baseline have been shown to predict increasing SPR concerns over time, but not PR concerns [Munro2022]. From a measurement perspective, dyadic CFI data may therefore reflect both the level of reported cognitive concerns and the relative balance between participant and study partner reports.

Despite these advances, it remains unclear whether Alzheimer’s disease biomarkers are associated primarily with higher levels of cognitive concerns across reporters, with reporter- specific imbalance, or with both features of dyadic CFI data. Longitudinal studies have shown that memory self-awareness discrepancy varies with amyloid burden [Vannini2017a; Hanseeuw2019], and self- and study partner-reported concerns have been associated with tau pathology [Jadick2024]. However, it remains unclear whether the regional extent of amyloid pathology is associated with the relative balance between participant- and study partner-reported CFI scores in cognitively unimpaired individuals. The regional extent of amyloid pathology [Ozlen2022; Farrell2024] may be more informative than global burden alone, particularly within early amyloid-vulnerable cortical regions, for capturing early awareness-related complaint patterns. Furthermore, the extent to which tau is associated with CFI scores across reporters or with reporter-specific imbalance remains poorly characterized. Finally, it is unknown whether tau PET and plasma p-tau217 [Janelidze2020; Palmqvist2020] provide similar information about these features of dyadic CFI data. This comparison is clinically relevant given the growing availability of blood-based biomarkers.

A prior study showed that both participant- and study partner-reported CFI ratings are associated with regional tau pathology across multiple cohorts [Jadick2024], but did not test pathology × reporter interactions across amyloid regional extent categories or directly compare tau PET with plasma p-tau217 within a unified dyadic model. The present study extends this work by distinguishing biomarker associations with CFI scores from biomarker associations with the relative balance between reporters, while also examining whether tau PET and plasma p- tau217 provide similar information within a unified dyadic model. The primary focus was therefore the biomarker-informed interpretation of dyadic CFI scores, rather than the presence or absence of group-level crossover between adjusted reporter means alone.

Accordingly, we examined whether amyloid regional extent and tau burden were associated with two features of dyadic CFI data: CFI scores across reporters and the relative balance between participant and study partner reports. Amyloid regional extent was the primary biomarker analysis. As secondary analyses, we examined plasma p-tau217 and compared the relative explanatory value of tau PET and plasma p-tau217 in participants with both biomarkers available. We hypothesized that, among individuals with CDR global score 0, biomarker associations could differ according to whether they reflected CFI burden across reporters or reporter-specific imbalance, without requiring group-level crossover of the adjusted reporter means.

## Methods

### Study design and participants

Data were drawn from two complementary studies: the Anti-Amyloid Treatment in Asymptomatic Alzheimer’s Disease (A4) randomized controlled trial examining solanezumab in individuals with preclinical AD [Sperling2023] and the Longitudinal Evaluation of Amyloid Risk and Neurodegeneration (LEARN) observational cohort [Sperling2024]. Both studies enrolled cognitively unimpaired older adults and collected identical assessments at screening.

The inclusion of LEARN alongside A4 preserves the lower-amyloid reference range [Sperling2020], enabling characterization of low and early amyloid burden. Plasma p-tau217 analyses were conducted in the subset with available blood biomarker data, which was derived from A4 participants in the present dataset. Participants were included if amyloid PET data were available at screening (florbetapir/AV45) and the baseline CDR global score was 0. Participants classified as A4 screen failures who did not enroll in LEARN, as well as participants with missing data required to define eligibility or covariates, were excluded.

### Biomarker Definitions

Amyloid burden was quantified using global florbetapir SUVR [Clark2012; Joshi2015]. For categorical analyses, we constructed A4/LEARN-adapted amyloid regional extent categories based on available regional florbetapir SUVR measures (Supplementary Methods and Table S1). Tau burden was assessed using flortaucipir PET. The primary continuous measure was an early-to-middle temporal tau composite, computed as the mean of z-scored bilateral entorhinal and inferior temporal cortex SUVR values [Schöll2016;Johnson2016;Sanchez2021]. Plasma p- tau217, measured at baseline, was standardized (z-scored) within all A4/LEARN participants with available blood data (N = 1,066) to allow comparability with other assays [Palmqvist2020].

### Outcomes and Covariates

The primary outcome was the reporter-specific Cognitive Function Index (CFI) total score, a 15- item dyadic measure of perceived cognitive change, with higher scores indicating greater perceived cognitive decline. Participant-reported and study partner-reported CFI observations were arranged in long format to estimate biomarker associations jointly across reporters and to test whether these associations differed between reporters. In the scored variables used for this analysis, reporter-specific CFI totals ranged from 0 to 15 for both participant report (CFI-PT) and study partner report (CFI-SP), with higher scores indicating greater perceived cognitive change. The combined total (CFI-Total, range 0–30) is the sum of both reports. Inspection of the A4/LEARN dataset confirmed that CFI-Total was set to missing whenever either reporter score was absent. The final analytic sample contained 1,682 participants. All models were adjusted for age, sex, years of education, APOE ε4 allele count (0, 1, or 2), and baseline cognition (PACC). Reporter-specific imbalance was defined as a differential biomarker association with CFI-PT versus CFI-SP and was indexed by the biomarker × reporter interaction term. Whether the adjusted mean CFI-SP score exceeded the adjusted mean CFI-PT score within a biomarker stratum was examined descriptively; this group-level crossover was not a primary estimand.

### Statistical analyses

To test whether amyloid burden differentially predicts participant- versus study partner-reported CFI, a long-format ordinary least squares (OLS) regression model was estimated with two observations per participant (CFI-PT and CFI-SP). For all long-format models, standard errors were clustered at the participant level using the sandwich estimator (vcovCL) to account for within-participant correlation between participant and study partner observations. The model was:

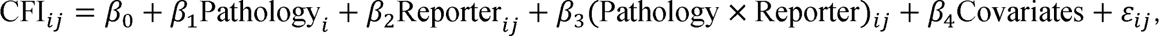

where *i* denotes the participant, *j* denotes the reporter, CFI-PT was the reporter reference, and *X_i_* included age, sex, years of education, *APOE* ε4 allele count, and PACC. For categorical biomarker models, category or stage 0 was used as the reference level.

The interaction term β_3_ (biomarker × reporter) constituted the primary estimand for reporter-specific imbalance and tested whether the biomarker–CFI association differed between CFI-PT and CFI-SP. Because interaction terms were prespecified primary estimands, p values are reported as nominal. For categorical amyloid regional extent and tau PET-based stage models, we additionally report Wald tests for the overall pathology × reporter interaction. Category- or stage-specific contrasts were interpreted as exploratory unless supported by the corresponding global interaction test. Findings outside the primary interaction framework were interpreted as supportive rather than definitive. A biomarker association without a detectable biomarker × reporter interaction was interpreted as an association with CFI scores that did not differ detectably between reporters, rather than as evidence of identical effects in the two reporters.

Detailed biomarker definitions, complementary analyses, nested model comparisons, and sensitivity analyses are described in the Supplementary Methods.

### Ethics

This study was approved by the University of Tokyo Graduate School of Medicine institutional ethics committee (ID: 2025264NI). The requirement for informed consent was waived because this secondary analysis used de-identified data obtained through an approved data access process.

## Results

### Sample characteristics

The combined A4/LEARN baseline sample included 1,686 cognitively unimpaired participants, of whom 1,682 had complete dyadic CFI data for the primary long-format models.

Characteristics according to the A4/LEARN-adapted regional amyloid patterns are shown in Table S2, including the number of participants per regional extent category, substudy composition, global amyloid SUVR, *APOE* ε4 allele count (0, 1, or 2), PACC, CFI-PT, CFI-SP, and ΔCFI. Because tau PET and plasma p-tau217 were available in selected subsamples, demographic and biomarker characteristics of the full sample, tau PET subsample, p-tau217 subsample, and matched tau PET/p-tau217 subsample are shown in Table S3. At baseline, 64.7% of participants were amyloid-positive (mean SUVR = 1.2 ± 0.2). Mean CFI scores were 2.2 ± 2.1 for participant report (CFI-PT) and 1.4 ± 1.9 for study partner report (CFI-SP), yielding a mean discordance score (ΔCFI = CFI-PT − CFI-SP) of 0.8 ± 2.2 (Table 1). Tau PET data were available for 438 participants, plasma p-tau217 for 1,066, and the matched subset with both biomarkers available comprised 352 participants (Figure S1). The plasma p-tau217 subset was entirely A4-derived and amyloid-enriched, whereas LEARN participants were not represented in the blood biomarker subset (Table S3).

**Table 1.**
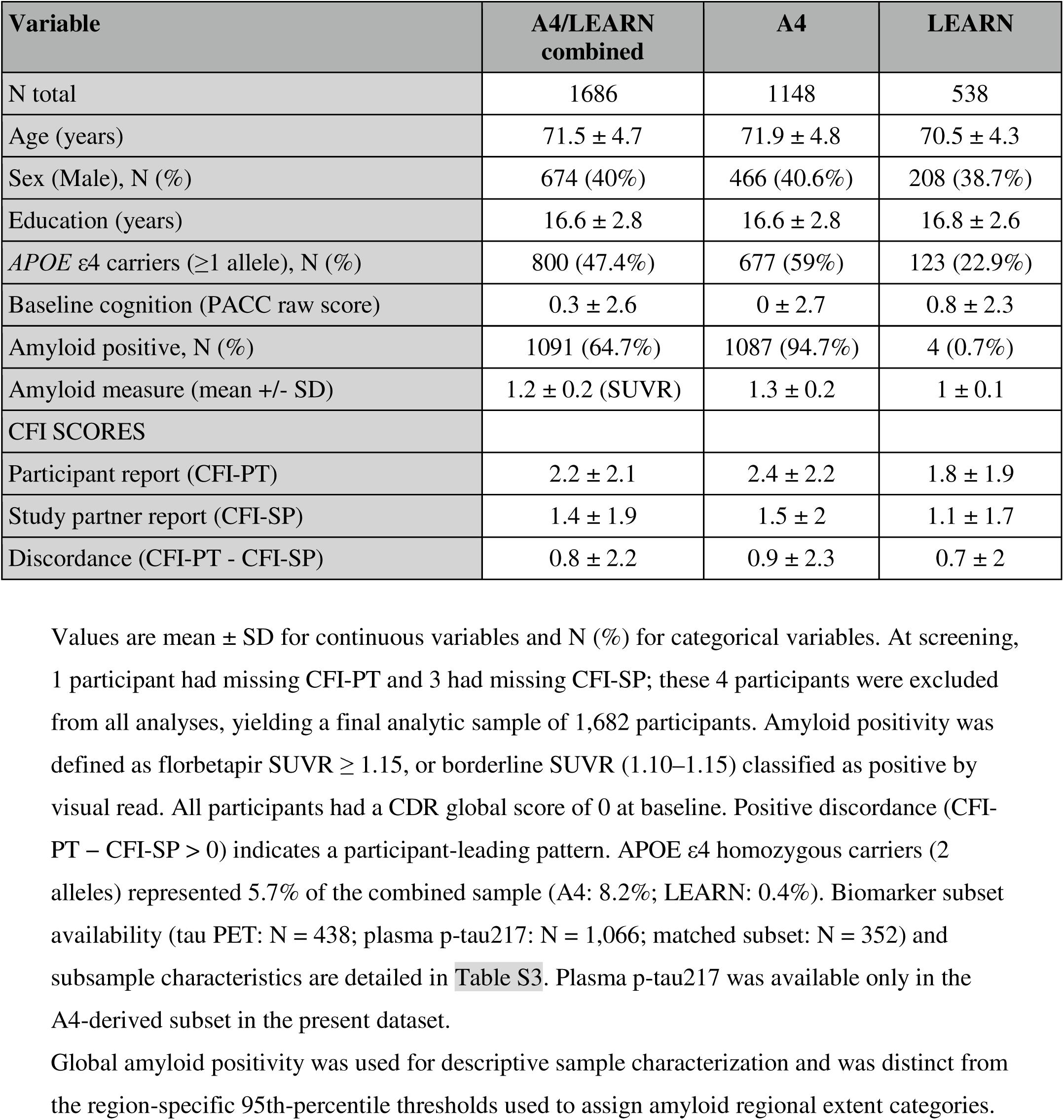
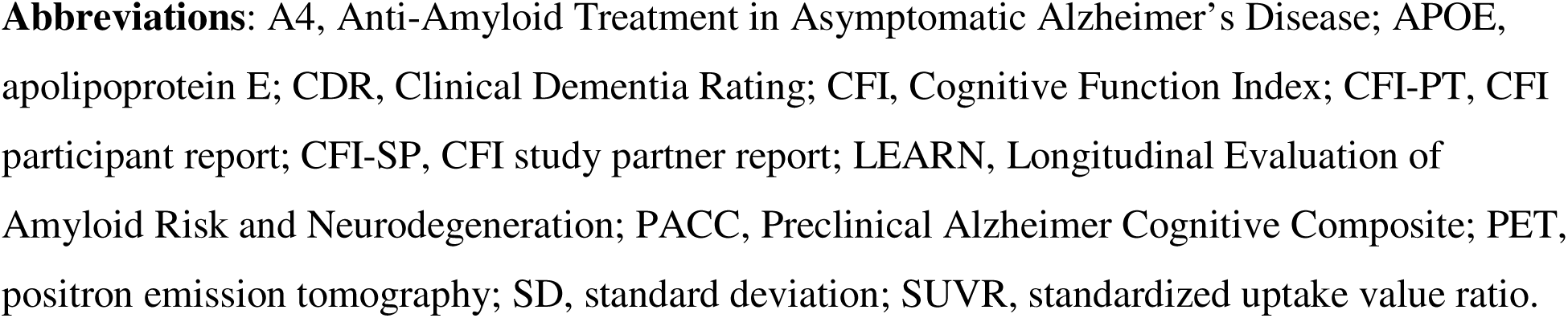
Baseline characteristics of the A4/LEARN screening sample. Values are mean ± SD for continuous variables and N (%) for categorical variables. At screening, 1 participant had missing CFI-PT and 3 had missing CFI-SP; these 4 participants were excluded from all analyses, yielding a final analytic sample of 1,682 participants. Amyloid positivity was defined as florbetapir SUVR ≥ 1.15, or borderline SUVR (1.10–1.15) classified as positive by visual read. All participants had a CDR global score of 0 at baseline. Positive discordance (CFI- PT − CFI-SP > 0) indicates a participant-leading pattern. APOE ε4 homozygous carriers (2 alleles) represented 5.7% of the combined sample (A4: 8.2%; LEARN: 0.4%). Biomarker subset availability (tau PET: N = 438; plasma p-tau217: N = 1,066; matched subset: N = 352) and subsample characteristics are detailed in Table S3. Plasma p-tau217 was available only in the A4-derived subset in the present dataset. Global amyloid positivity was used for descriptive sample characterization and was distinct from the region-specific 95th-percentile thresholds used to assign amyloid regional extent categories. **Abbreviations**: A4, Anti-Amyloid Treatment in Asymptomatic Alzheimer’s Disease; APOE, apolipoprotein E; CDR, Clinical Dementia Rating; CFI, Cognitive Function Index; CFI-PT, CFI participant report; CFI-SP, CFI study partner report; LEARN, Longitudinal Evaluation of Amyloid Risk and Neurodegeneration; PACC, Preclinical Alzheimer Cognitive Composite; PET, positron emission tomography; SD, standard deviation; SUVR, standardized uptake value ratio.

### Amyloid Regional Extent and Reporter Balance

In the long-format amyloid model, the overall amyloid regional extent category × reporter interaction was significant (Wald test, df = 3, p = 0.0016), indicating that the balance between CFI-PT and CFI-SP differed across regional extent categories (Table 2). Amyloid regional extent categories were also associated with higher CFI scores in the reference reporter, CFI-PT, and study partner report was consistently lower than participant report (β = −0.641, p < 0.001).

**Table 2.**
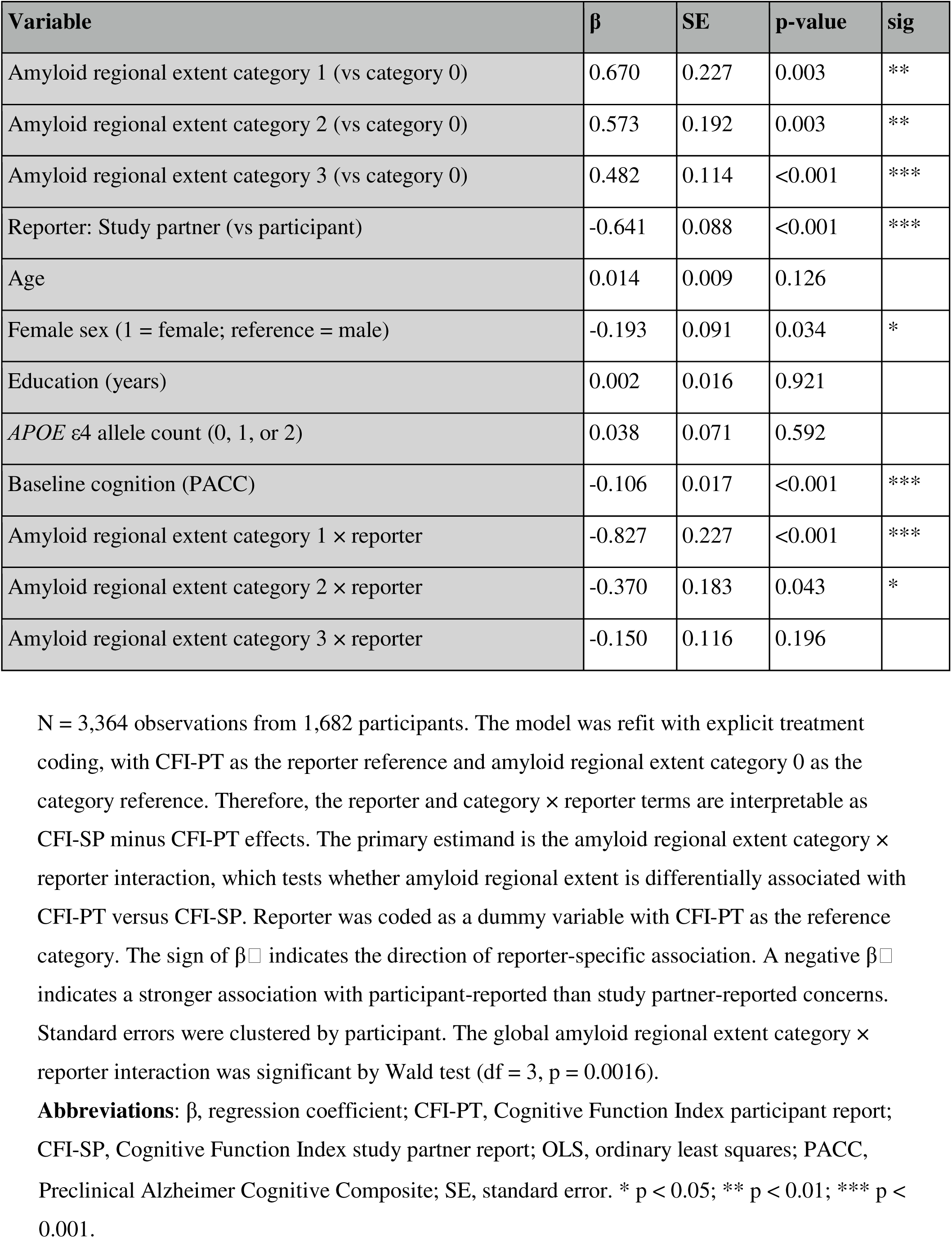
Amyloid regional extent and reporter balance: long-format interaction model. N = 3,364 observations from 1,682 participants. The model was refit with explicit treatment coding, with CFI-PT as the reporter reference and amyloid regional extent category 0 as the category reference. Therefore, the reporter and category × reporter terms are interpretable as CFI-SP minus CFI-PT effects. The primary estimand is the amyloid regional extent category × reporter interaction, which tests whether amyloid regional extent is differentially associated with CFI-PT versus CFI-SP. Reporter was coded as a dummy variable with CFI-PT as the reference category. The sign of β indicates the direction of reporter-specific association. A negative β indicates a stronger association with participant-reported than study partner-reported concerns. Standard errors were clustered by participant. The global amyloid regional extent category × reporter interaction was significant by Wald test (df = 3, p = 0.0016). **Abbreviations**: β, regression coefficient; CFI-PT, Cognitive Function Index participant report; CFI-SP, Cognitive Function Index study partner report; OLS, ordinary least squares; PACC, Preclinical Alzheimer Cognitive Composite; SE, standard error. * p < 0.05; ** p < 0.01; *** p < 0.001.

Category-specific contrasts showed the strongest participant-leading interaction for the restricted early cortical amyloid pattern (category 1: β_3_ = −0.827, p < 0.001). A weaker nominal interaction was observed for parietal and/or anterior cingulate involvement without temporal/diffuse involvement (category 2: β_3_ = −0.370, p = 0.043), whereas the temporal/diffuse pattern (category 3) did not show a significant interaction. Adjusted reporter-specific means across amyloid regional extent categories are shown in Figure 1A. Thus, the global interaction was driven primarily by the large participant-leading contrast in the restricted early cortical subgroup, although this category-specific finding was exploratory. It should be noted that amyloid regional extent category 1 was a relatively small subgroup (N = 87), with distinct substudy composition. Separate-reporter models showed consistent directionality (Supplementary Results, Table S4). A complementary continuous amyloid model and direct discrepancy analyses are provided in Tables S5 and S6, respectively. Notably, the category 1 association remained significant in the direct ΔCFI model, in which sex and the other covariates were modeled as predictors of the reporter difference (β = 0.728, p = 0.004; Table S6).

**Figure 1.**
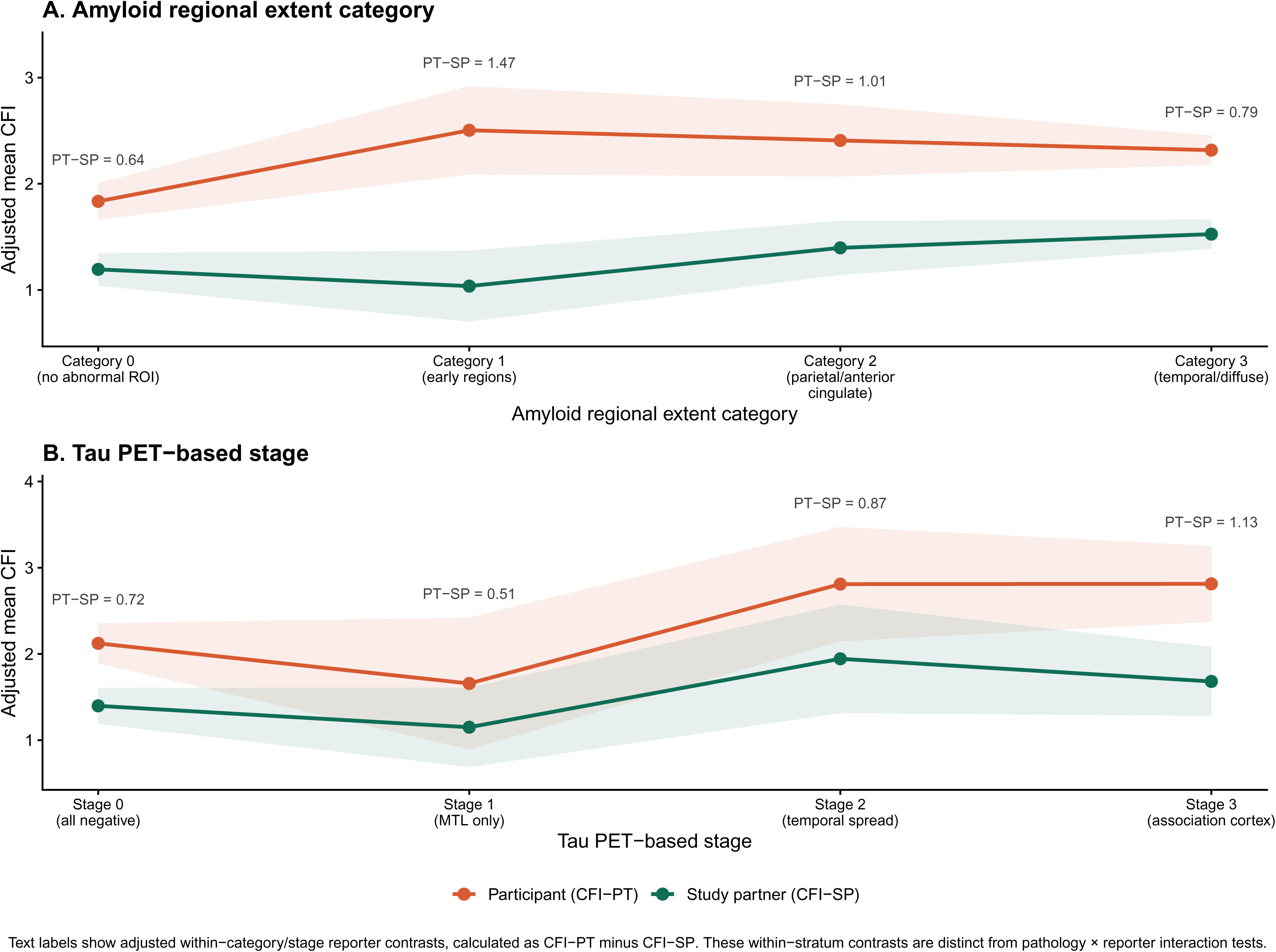
Adjusted mean CFI scores by reporter across amyloid regional extent categories and tau PET-based stages. Adjusted estimated marginal means and 95% confidence intervals were derived from the corresponding long-format linear models. Panel A shows adjusted CFI-PT and CFI-SP scores across amyloid regional extent categories in the full dyadic CFI sample (N=1,682). Reporter balance varied across amyloid regional extent categories, with the largest adjusted participant- leading contrast observed in category 1. Formal inference was based on the global category × reporter interaction and the corresponding category-specific contrasts. Panel B shows adjusted CFI-PT and CFI-SP scores across tau PET-based stages in the tau PET subsample. These descriptive estimates complement the primary continuous tau PET model, in which the tau PET association with CFI did not differ detectably between reporters. Text labels indicate the adjusted within-category or within-stage reporter contrast, calculated as CFI-PT minus CFI-SP. These within-stratum contrasts describe the magnitude of participant-leading reporting within each category or stage and are distinct from the pathology × reporter interaction terms used as the primary inferential tests. All models were adjusted for age, sex, years of education, *APOE* ε4 allele count, and PACC; the tau model was additionally adjusted for amyloid regional extent category. **Abbreviations**: APOE, apolipoprotein E; CFI-PT, Cognitive Function Index participant report; CFI-SP, Cognitive Function Index study partner report; CI, confidence interval; PACC, Preclinical Alzheimer Cognitive Composite; PET, positron emission tomography. Adjusted mean CFI scores by reporter across amyloid regional extent categories and tau PET−based stages

### Tau PET Associations With CFI Scores Across Reporters

The early-to-middle temporal tau composite was associated with higher CFI scores across reporters (β = 0.334, SE = 0.112, p = 0.003; Table 3). Descriptive stage-based analyses and adjusted reporter-specific means are provided in the Supplementary Results, Table S7, and Figure 1B. Adding the tau composite as a main effect improved model fit beyond the amyloid-only model (p < 0.001; Table S8). The temporal tau composite × reporter interaction was not significant (β = 0.035, SE = 0.151, p = 0.816; approximate 95% CI, −0.263 to 0.333; Table 3), and adding this interaction did not improve model fit (p = 0.817; Table S8). These results indicate that tau PET was associated with higher CFI scores, but the magnitude of this association did not differ detectably between participant and study partner reports.

**Table 3.** Tau PET associations with CFI scores: main-effect model and reporter interaction test. All rows except the final row are from the tau main-effect model. The final row shows the additional tau × reporter term from the nested interaction model. Other coefficients from the nested interaction model are not displayed. The tau × reporter interaction tests whether the association between tau burden and CFI differs between CFI-PT and CFI-SP. Standard errors were clustered by participant. Results were estimated in the tau PET subsample (N = 438 participants; 876 long-format observations). Amyloid regional extent coefficients in this table are covariate estimates within the selected tau PET subsample and should not be interpreted as the primary amyloid estimates, which are reported in Table 2. **Abbreviations**: β, regression coefficient; CFI-PT, Cognitive Function Index participant report; CFI-SP, Cognitive Function Index study partner report; PACC, Preclinical Alzheimer Cognitive Composite; PET, positron emission tomography; SE, standard error; * p < 0.05; ** p < 0.01; *** p < 0.001.

| Variable | $\beta$ | SE | p-value | sig |
| --- | --- | --- | --- | --- |
| (Intercept) | 0.920 | 1.491 | 0.537 |  |
| Amyloid regional extent category 1 | 0.777 | 0.488 | 0.111 |  |
| Amyloid regional extent category 2 | 0.009 | 0.413 | 0.982 |  |
| Amyloid regional extent category 3 | -0.146 | 0.306 | 0.634 |  |
| Reporter: study partner (vs participant) | -0.578 | 0.293 | 0.049 | * |
| Early-to-middle temporal tau composite | 0.334 | 0.112 | 0.003 | ** |
| Age | 0.014 | 0.019 | 0.468 |  |
| Female sex (1 = female; reference = male) | -0.295 | 0.166 | 0.077 |  |
| Education (years) | 0.037 | 0.030 | 0.209 |  |
| APOE ε4 allele count (0, 1, or 2) | 0.128 | 0.137 | 0.351 | |
| Baseline cognition (PACC) | -0.106 | 0.029 | <0.001 | *** |
| Amyloid category 1 × study partner | -1.672 | 0.526 | 0.002 | ** |
| Amyloid category 2 × study partner | -0.526 | 0.402 | 0.190 | |
| Amyloid category 3 × study partner | -0.165 | 0.326 | 0.613 | |
| Study partner × early-to-middle temporal tau composite | 0.035 | 0.151 | 0.816 | |

### Plasma p-tau217 Associations With Dyadic CFI Scores

Plasma p-tau217 was available in an A4-derived, amyloid-enriched subset of 1,066 participants; the complete-case long-format model included 1,065 participants. In the A4-derived subset with available plasma p-tau217 (Table 4), the estimated association between higher z-scored plasma p-tau217 and overall CFI scores was positive but did not meet the conventional significance threshold (β = 0.149, SE = 0.077, p = 0.053). The p-tau217 × reporter interaction was not significant (p = 0.810). Thus, this selected subset provided no clear evidence of either a p-tau217 association with CFI scores at the conventional significance threshold or a reporter-specific p- tau217 association. Study partner report remained lower than participant report (β = −0.854, p < 0.001).

**Table 4.** Plasma p-tau217 associations with CFI scores and reporter-specific imbalance. N = 2,130 observations from 1,065 participants after listwise deletion. Plasma p-tau217 was available for 1,066 participants; one participant was excluded from this model because of missing CFI or covariate data. The model was refit with explicit treatment coding, with CFI-PT as the reporter reference. Therefore, the reporter and p-tau217 × reporter terms are interpretable as CFI-SP minus CFI-PT effects. The primary estimand is the p-tau217 × reporter interaction, which tests whether plasma p-tau217 is differentially associated with CFI-PT versus CFI-SP. All models were adjusted for amyloid regional extent category, age, sex, years of education, APOE ε4 allele count, and baseline cognition (PACC). Standard errors were clustered by participant. **Abbreviations**: β, regression coefficient; CFI-PT, Cognitive Function Index participant report; CFI-SP, Cognitive Function Index study partner report; PACC, Preclinical Alzheimer Cognitive Composite; p-tau217, phosphorylated tau 217; SE, standard error. *** p < 0.001; . p < 0.10.

| Variable | $\beta$ | SE | p-value | sig |
| --- | --- | --- | --- | --- |
| (Intercept) | 0.762 | 1.018 | 0.455 |  |
| Reporter: study partner (vs participant) | -0.854 | 0.070 | <0.001 | *** |
| p-tau217 (z-score) | 0.149 | 0.077 | 0.053 | . |
| Amyloid regional extent category 1 (vs category 0) | 0.042 | 0.250 | 0.866 |  |
| Amyloid regional extent category 2 (vs category 0) | 0.263 | 0.238 | 0.269 |  |
| Amyloid regional extent category 3 (vs category 0) | 0.251 | 0.203 | 0.218 |  |
| Age | 0.019 | 0.012 | 0.135 |  |
| Female sex (1 = female; reference = male) | -0.197 | 0.118 | 0.094 | . |
| Education (years) | 0.005 | 0.021 | 0.810 |  |
| APOE ε4 allele count (0, 1, or 2) | 0.038 | 0.086 | 0.655 | |
| Baseline cognition (PACC) | -0.127 | 0.022 | <0.001 | *** |
| Study partner report × p-tau217 | -0.019 | 0.079 | 0.810 | |

### Secondary Comparison of Tau PET and Plasma p-tau217

Within the matched subsample and unified long-format analytical framework, the model including tau PET showed better fit than the baseline amyloid regional extent × reporter model and the corresponding plasma p-tau217 model (AIC = 2,983.6; adjusted R² = 0.105; Table 5). Tau PET added significant information beyond p-tau217 (p = 0.007), whereas p-tau217 did not add beyond tau PET (p = 0.527). The model-fit advantage of tau PET over plasma p-tau217 was not attributable to a reporter-specific association, as neither biomarker showed a significant biomarker × reporter interaction. This comparison concerned cross-sectional CFI model fit and should not be interpreted as a comparison of diagnostic or prognostic performance.

**Table 5.**
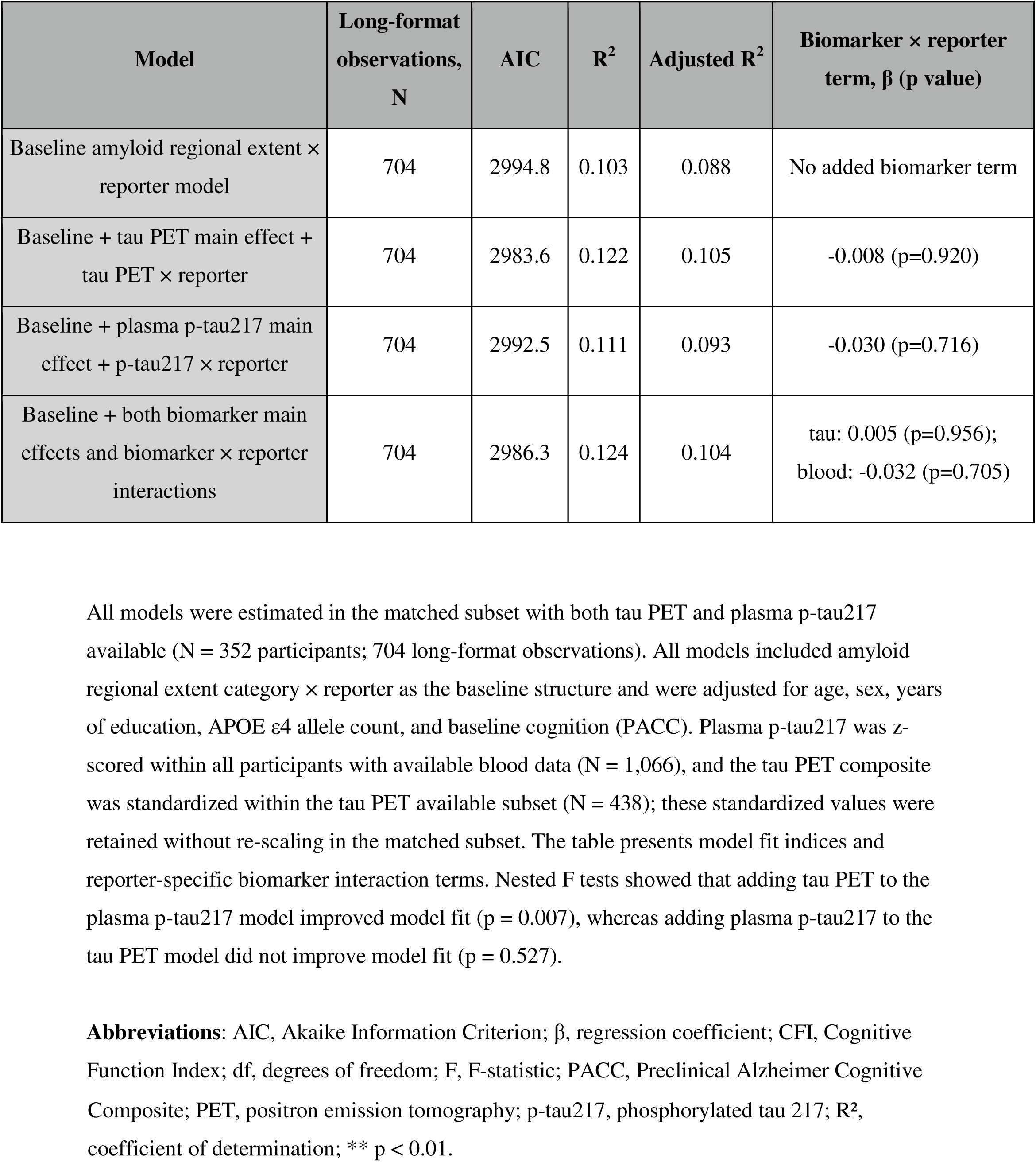
Secondary comparison of tau PET and plasma p-tau217: model fit indices. All models were estimated in the matched subset with both tau PET and plasma p-tau217 available (N = 352 participants; 704 long-format observations). All models included amyloid regional extent category × reporter as the baseline structure and were adjusted for age, sex, years of education, APOE ε4 allele count, and baseline cognition (PACC). Plasma p-tau217 was z- scored within all participants with available blood data (N = 1,066), and the tau PET composite was standardized within the tau PET available subset (N = 438); these standardized values were retained without re-scaling in the matched subset. The table presents model fit indices and reporter-specific biomarker interaction terms. Nested F tests showed that adding tau PET to the plasma p-tau217 model improved model fit (p = 0.007), whereas adding plasma p-tau217 to the tau PET model did not improve model fit (p = 0.527). **Abbreviations**: AIC, Akaike Information Criterion; β, regression coefficient; CFI, Cognitive Function Index; df, degrees of freedom; F, F-statistic; PACC, Preclinical Alzheimer Cognitive Composite; PET, positron emission tomography; p-tau217, phosphorylated tau 217; R^2^, coefficient of determination; ** p < 0.01.

## Discussion

The present study used a joint dyadic model to distinguish biomarker associations with CFI scores from biomarker associations with the relative balance between participant and study partner reports. The findings suggest that these two features of dyadic CFI data may have partly different biomarker correlates. Amyloid regional extent modified reporter balance, with the largest participant-leading contrast in the restricted early cortical subgroup. By contrast, temporal tau PET was associated with higher CFI scores without a detectable reporter-specific difference, whereas plasma p-tau217 showed no clear association in the selected A4-derived subset. These cross-sectional findings describe group-level associations and should not be interpreted as within-person awareness changes.

Within the range of CDR global score 0, adjusted mean CFI-SP did not exceed adjusted mean CFI-PT in any biomarker stratum. Thus, the group-level reporter means did not show a pathology-associated inversion, although the magnitude of the participant-leading difference varied across amyloid regional extent categories. This is consistent with evidence that frank anosognosia is more clearly observed once cognitive impairment reaches functional impact, such as at CDR ≥ 0.5 [Vannini2017a;Gagliardi2021]. Despite this absence of frank crossover, the pathology × reporter interaction model revealed a significant and category-dependent pattern: restricted early cortical amyloid abnormality was associated with maximal participant-leading discordance, with a similar but weaker pattern in the parietal/anterior cingulate pattern and attenuation in the temporal/diffuse pattern. This participant-leading profile is consistent with prior descriptions of heightened self-awareness in amyloid-positive cognitively normal individuals, but reporter imbalance is an indirect measure and the present cross-sectional analysis does not establish heightened awareness or a distinct hypernosognosia phase at the individual level. This phenomenon is closely related to subjective cognitive decline [Jessen2014;Cacciamani2026]. The attenuation of reporter divergence in category 3 appeared to reflect higher study partner reporting rather than lower participant reporting. This pattern is compatible with prior longitudinal evidence that study partner concerns become more closely aligned with pathological burden over time [Munro2022].

The present findings extend prior work by distinguishing biomarker associations with CFI scores from biomarker associations with reporter balance. Prior work suggests that awareness of cognitive change may follow several patterns across CN and MCI populations [Vannini2017a], and that amyloid pathology is associated with altered memory self-awareness [Hanseeuw2019]. Whereas prior work examined associations between regional tau and participant- and study partner-reported CFI ratings across multiple cohorts [Jadick2024], the present study jointly modeled both reporters to distinguish associations with CFI scores from reporter-specific imbalance and examined whether tau PET and plasma p-tau217 provided similar information. Consistent with the present findings, previous work showed that both participant and study partner CFI increased over the course of the A4 Study, but that study partner CFI became more strongly aligned with objective cognitive decline over time, particularly among those in the highest amyloid tertile [Amariglio2024]. This longitudinal pattern is consistent with the cross-sectional reporter imbalance observed here, in which study partner report became clearly elevated only in the broader amyloid involvement categories, supporting the view that the relative importance of participant- and study partner-reported concerns may shift with increasing amyloid burden. The present findings are also consistent with the broader subjective cognitive decline literature [Jessen2014], which has shown that self-reported cognitive concerns in cognitively normal individuals are associated with amyloid pathology and may precede objective decline [Vannini2017a; Munro2022; Cacciamani2026].

The early-to-middle temporal tau PET composite added significant explanatory power beyond amyloid for overall CFI burden, but showed no significant tau × reporter interaction. This indicates that, in this cognitively unimpaired tau PET subsample, temporal tau burden was associated with CFI scores without evidence that the association differed between participant and study partner reports. The estimated plasma p-tau217 association with overall CFI scores was positive but did not meet the conventional significance threshold. In the head-to-head comparison, tau PET added significant independent information beyond p-tau217 but not the reverse, although the absolute difference in explained variance was small. The incremental model-fit contribution of tau PET reflected overall CFI burden rather than a reporter-specific signal, as neither biomarker showed a significant biomarker × reporter interaction. The head-to-head analysis was designed to compare explanatory information for cross-sectional CFI scores and should not be interpreted as a comparison of the diagnostic or prognostic performance of tau PET and plasma p-tau217.

Several limitations should be acknowledged. First, the cross-sectional design precludes inference about within-person awareness trajectories or phase transitions. Second, CFI discordance is an indirect proxy for awareness-related complaint patterns and may be influenced by mood, anxiety, personality, study partner relationship, and contact frequency, which were not fully modeled here. Third, the amyloid regional extent categories were adapted to the limited regional florbetapir SUVR measures available in A4/LEARN and should not be interpreted as a direct implementation of validated amyloid PET staging systems. Fourth, A4 and LEARN differed substantially in amyloid enrichment and biomarker availability, and amyloid regional extent category and substudy composition cannot be fully separated in this dataset. The restricted early cortical amyloid group was relatively small (N = 87) and had a substantially different A4/LEARN composition from the other regional extent categories. Therefore, the category-specific participant-leading finding should be considered a hypothesis-generating cross-sectional result and requires replication in independent cohorts with richer regional amyloid measures.

Fifth, tau PET and plasma p-tau217 were available in selected subsamples enriched for A4 participants and amyloid-positive individuals, limiting generalizability. Sixth, the A4/LEARN cohort is a trial-screening sample that is highly educated and relatively homogeneous; findings may not generalize to more diverse or population-based samples, although prior work has demonstrated measurement invariance of the CFI across racial/ethnic groups within A4 [Ruthirakuhan2024].

In conclusion, joint interpretation of participant- and study partner-reported CFI scores may help distinguish biomarker associations with CFI scores from biomarker associations involving reporter-specific imbalance. Amyloid regional extent was associated with reporter balance, whereas tau PET was associated with CFI scores without detectable reporter-specific divergence. These cross-sectional findings require independent replication and longitudinal validation.

## Data Availability

Data used in this study were obtained through the Alzheimer's Clinical Trial Consortium data access process. Qualified investigators may request access to A4/LEARN data through the ACTC data access platform (https://www.actcinfo.org), subject to applicable data-use requirements.

## Acknowledgements

The authors acknowledge the A4 and LEARN Study Teams for their important contributions to the A4 and LEARN Studies. The A4 Study was funded by a public-private-philanthropic partnership, including the National Institutes of Health/National Institute on Aging (R01AG063689, U19AG010483, and U24AG057437), Eli Lilly and Company, the Alzheimer’s Association, the Accelerating Medicines Partnership through the Foundation for the National Institutes of Health, the GHR Foundation, the Davis Alzheimer Prevention Program, an anonymous foundation, and additional private donors to Brigham and Women’s Hospital, with in-kind support from Avid, Cogstate, Albert Einstein College of Medicine, and the Foundation for Neurologic Diseases. The companion observational Longitudinal Evaluation of Amyloid Risk and Neurodegeneration (LEARN) Study was funded by the Alzheimer’s Association and the GHR Foundation. The complete A4 and LEARN Study Team list is available through the ACTC A4 Study Team Lists page.

The authors’ affiliation, “*Dementia Inclusion and Therapeutics*,” is an endowed department funded by Effissimo Capital Management Pte Ltd. AI-assisted tools were used for language editing; authors take full responsibility for the content.

## Consent Statement

The requirement for informed consent for this secondary analysis was waived by the institutional ethics committee because only de-identified data obtained through an approved data access process were used.

## References

1. Caselli RJ, Reiman EM. Characterizing the Preclinical Stages of Alzheimer’s Disease and the Prospect of Presymptomatic Intervention. J Alzheimer’s Dis. 2013 Jan 1;33(s1):S405–16. 10.3233/JAD-2012-129026. PubMed PMID: 22695623.

2. Sperling RA, Aisen PS, Beckett LA, Bennett DA, Craft S, Fagan AM, Iwatsubo T, Jack CR Jr, Kaye J, Montine TJ, Park DC, Reiman EM, Rowe CC, Siemers E, Stern Y, Yaffe K, Carrillo MC, Thies B, Morrison-Bogorad M, Wagster MV, Phelps CH. Toward defining the preclinical stages of Alzheimer’s disease: recommendations from the National Institute on Aging-Alzheimer’s Association workgroups on diagnostic guidelines for Alzheimer’s disease. Alzheimers Dement. 2011 May;7(3):280–92. 10.1016/j.jalz.2011.03.003. Epub 2011 Apr 21. PubMed PMID: 21514248.

3. Vannini P, Amariglio R, Hanseeuw B, et al. Memory self-awareness in the preclinical and prodromal stages of Alzheimer’s disease. Neuropsychologia. 2017;99:343–349. PMID: 28385579. DOI: 10.1016/j.neuropsychologia.2017.04.002.

4. Reed BR, Jagust WJ, Coulter L. Anosognosia in Alzheimer’s disease: Relationships to depression, cognitive function, and cerebral perfusion. J Clin Exp Neuropsychol. 1993 Mar 1;15(2):231–44. 10.1080/01688639308402560. PubMed PMID: 8491848.

5. Cacciamani F, Houot M, Gagliardi G, Dubois B, Sikkes S, Sánchez-Benavides G, et al. Awareness of Cognitive Decline in Patients With Alzheimer’s Disease: A Systematic Review and Meta-Analysis. Front Aging Neurosci. 2021 Aug 3;13. 10.3389/fnagi.2021.697234. PubMed PMID: 34413767.

6. Vannini P, Hanseeuw B, Munro CE, Amariglio RE, Marshall GA, Rentz DM, et al. Anosognosia for memory deficits in mild cognitive impairment: Insight into the neural mechanism using functional and molecular imaging. NeuroImage Clin. 2017 Jan 1;15:408–14. 10.1016/j.nicl.2017.05.020. PubMed PMID: 28616381.

7. Therriault J, Ng KP, Pascoal TA, Mathotaarachchi S, Kang MS, Struyfs H, et al. Anosognosia predicts default mode network hypometabolism and clinical progression to dementia. Neurology. 2018 Mar 13;90(11):e932–9. 10.1212/WNL.0000000000005120. PubMed PMID: 29444971.

8. Hanseeuw BJ, Scott MR, Sikkes SAM, Properzi M, Gatchel JR, Salmon E, et al. Evolution of anosognosia in alzheimer’s disease and its relationship to amyloid. Ann Neurol. 2020;87(2):267–80. 10.1002/ana.25649. PubMed PMID: 31750553.

9. Walsh SP, Raman R, Jones KB, Aisen PS, Group for the ADCS. ADCS Prevention Instrument Project: The Mail-In Cognitive Function Screening Instrument (MCFSI). Alzheimer Dis Assoc Disord. 2006 Dec;20: S170–S178 10.1097/01.wad.0000213879.55547.57. PubMed PMID: 17135810.

10. Amariglio RE, Donohue MC, Marshall GA, Rentz DM, Salmon DP, Ferris SH, Karantzoulis S, Aisen PS, Sperling RA. Tracking early decline in cognitive function in older individuals at risk for Alzheimer disease dementia. JAMA Neurol. 2015;72(4):446. 10.1001/jamaneurol.2014.3375. PubMed PMID: 25706191.

11. Buckley R, Saling M, Ellis K, et al. Self and informant memory concerns align in healthy memory complainers and in early stages of mild cognitive impairment but separate with increasing cognitive impairment. Age Ageing. 2015;44(6):1012–1019. PMID: 26452663. DOI: 10.1093/ageing/afv136.

12. Amariglio R, Chou HCL, Buckley RF, et al. The dynamic interplay between longitudinal subjective and objective cognitive decline along the early AD spectrum in the Harvard Aging Brain Study. Alzheimers Dement. 2020;16(S6):e040260. DOI: 10.1002/alz.040260.

13. Munro CE, Buckley R, Vannini P, DeMuro C, Sperling R, Rentz DM, et al. Longitudinal Trajectories of Participant- and Study Partner-Rated Cognitive Decline, in Relation to Alzheimer’s Disease Biomarkers and Mood Symptoms. Front Aging Neurosci. 2022 Jan 31;13. 10.3389/fnagi.2021.806432. PubMed PMID: 35173601.

14. Hanseeuw BJ, Betensky RA, Jacobs HIL, Schultz AP, Sepulcre J, Becker JA, et al. Association of Amyloid and Tau With Cognition in Preclinical Alzheimer Disease: A Longitudinal Study. JAMA Neurol. 2019 Aug 1;76(8):915–24. 10.1001/jamaneurol.2019.1424 . PubMed PMID: 31157827.

15. Jadick MF, Robinson T, Farrell ME, Klinger H, Buckley RF, Marshall GA, et al. Associations Between Self and Study Partner Report of Cognitive Decline With Regional Tau in a Multicohort Study. Neurology. 2024 Jun 25;102(12):e209447. 10.1212/WNL.0000000000209447. PubMed PMID: 38810211

16. Ozlen H, Binette AP, Köbe T, Meyer P, Gonneaud J, St-Onge F, et al. Spatial extent of amyloid-β levels and associations with tau-PET and cognition. JAMA Neurol. 2022;79(10):1025. 10.1001/jamaneurol.2022.2442. PubMed PMID: 35994280.

17. Farrell ME, Thibault EG, Becker JA, Price JC, Healy BC, Hanseeuw BJ, Buckley RF, Jacobs HIL, Schultz AP, Chen CD, Sperling RA, Johnson KA. Spatial extent as a sensitive amyloid-PET metric in preclinical Alzheimer’s disease. Alzheimers Dement. 2024;20(8):5434–5449. 10.1002/alz.14036. PubMed PMID: 38988055.

18. Janelidze S, Stomrud E, Smith R, Palmqvist S, Mattsson N, Airey DC, et al. Cerebrospinal fluid p-tau217 performs better than p-tau181 as a biomarker of Alzheimer’s disease. Nat Commun. 2020 Apr 3;11(1):1683. 10.1038/s41467-020-15436-0. PubMed PMID: 32246036

19. Palmqvist S, Janelidze S, Quiroz YT, et al. Discriminative accuracy of plasma phospho- tau217 for Alzheimer disease vs other neurodegenerative disorders. JAMA. 2020;324(8):772–781. PMID: 32722745. DOI: 10.1001/jama.2020.12134.

20. Sperling RA, Donohue MC, Raman R, Rafii MS, Johnson K, Masters CL, et al. Trial of Solanezumab in Preclinical Alzheimer’s Disease. N Engl J Med. 2023 Sep 20;389(12):1096–107. 10.1056/nejmoa2305032. PubMed PMID: 37458272

21. Sperling RA, Donohue MC, Rissman RA, Johnson KA, Rentz DM, Grill JD, et al. Amyloid and Tau Prediction of Cognitive and Functional Decline in Unimpaired Older Individuals: Longitudinal Data from the A4 and LEARN Studies. J Prev Alzheimers Dis. 2024 Aug 1;11(4):802–13. 10.14283/jpad.2024.122. PubMed PMID: 39044488

22. Sperling RA, Donohue MC, Raman R, Sun CK, Yaari R, Holdridge K, et al. Association of Factors With Elevated Amyloid Burden in Clinically Normal Older Individuals. JAMA Neurol. 2020 Jun 1;77(6):735–45. 10.1001/jamaneurol.2020.0387. PubMed PMID: 32250387

23. Clark CM, Pontecorvo MJ, Beach TG, Bedell BJ, Coleman RE, Doraiswamy PM, et al. Cerebral PET with florbetapir compared with neuropathology at autopsy for detection of neuritic amyloid-β plaques: a prospective cohort study. Lancet Neurol. 2012 Aug;11(8):669–78. 10.1016/S1474-4422(12)70142-4. PubMed PMID: 22749065.

24. Joshi AD, Pontecorvo MJ, Lu M, Skovronsky DM, Mintun MA, Devous MD. A Semiautomated Method for Quantification of F 18 Florbetapir PET Images. J Nucl Med Off Publ Soc Nucl Med. 2015 Nov;56(11):1736–41. 10.2967/jnumed.114.153494. PubMed PMID: 26338898.

25. Schöll M, Lockhart SN, Schonhaut DR, O’Neil JP, Janabi M, Ossenkoppele R, et al. PET Imaging of Tau Deposition in the Aging Human Brain. Neuron. 2016 Mar 2;89(5):971–82. 10.1016/j.neuron.2016.01.028. PubMed PMID: 26938442.

26. Johnson KA, Schultz A, Betensky RA, Becker JA, Sepulcre J, Rentz D, et al. Tau positron emission tomographic imaging in aging and early Alzheimer disease. Ann Neurol. 2016 Jan;79(1):110–9. 10.1002/ana.24546. PubMed PMID: 26505746.

27. Sanchez JS, Becker JA, Jacobs HIL, Hanseeuw BJ, Jiang S, Schultz AP, et al. The cortical origin and initial spread of medial temporal tauopathy in Alzheimer’s disease assessed with positron emission tomography. Sci Transl Med. 2021 Jan 20;13(577):eabc0655. 10.1126/scitranslmed.abc0655. PubMed PMID: 33472953.

28. Gagliardi G, Kuppe M, Lois C, Hanseeuw B, Vannini P, for the Alzheimer’s Disease Neuroimaging Initiative. Pathological correlates of impaired self-awareness of memory function in Alzheimer’s disease. Alzheimers Res Ther. 2021 Jun 25;13(1):118. 10.1186/s13195-021-00856-x. PubMed PMID: 34172086

29. Jessen F, Amariglio RE, van Boxtel M, Breteler M, Ceccaldi M, Chételat G, Dubois B, Dufouil C, Ellis KA, van der Flier WM, Glodzik L, van Harten AC, de Leon MJ, McHugh P, Mielke MM, Molinuevo JL, Mosconi L, Osorio RS, Perrotin A, Petersen RC, Rabin LA, Rami L, Reisberg B, Rentz DM, Sachdev PS, de la Sayette V, Saykin AJ, Scheltens P, Shulman MB, Slavin MJ, Sperling RA, Stewart R, Uspenskaya O, Vellas B, Visser PJ, Wagner M; Subjective Cognitive Decline Initiative (SCD-I) Working Group. A conceptual framework for research on subjective cognitive decline in preclinical Alzheimer’s disease. Alzheimers Dement. 2014 Nov;10(6):844–52. 10.1016/j.jalz.2014.01.001. Epub 2014 May 3. PMID: 24798886.

30. Cacciamani F, Houot M, Tezenas du Montcel S, Thibeau-Sutre E, Vannini P, Migliaccio RL. Multimodal neural correlates of cognitive awareness in aging and Alzheimer’s disease. Neuroimage Clin. 2026;49:103922. PMID: 41380294. DOI: 10.1016/j.nicl.2025.103922.

31. Amariglio RE, Grill J, Rentz D, Marshall G, Donohue M, Liu A, Aisen P, Sperling R. Longitudinal trajectories of the Cognitive Function Index in the A4 Study. J Prev Alzheimers Dis. 2024;11(4):838–845. 10.14283/jpad.2024.125. PubMed PMID: 39044492.

32. Ruthirakuhan M, Alexander MW, Cogo-Moreira H, et al. Investigating the factor structure of the Preclinical Alzheimer Cognitive Composite and Cognitive Function Index across racial/ethnic, sex, and Aβ status groups in the A4 Study. J Prev Alzheimers Dis. 2024;11(1):48–55. PMID: 38230716. DOI: 10.14283/jpad.2023.98.

33. Mattsson N, Palmqvist S, Stomrud E, Vogel J, Hansson O. Staging β-Amyloid Pathology With Amyloid Positron Emission Tomography. JAMA Neurol. 2019 Nov 1;76(11):1319–29. 10.1001/jamaneurol.2019.2214. PubMed PMID: 31314895.

34. Grothe MJ, Barthel H, Sepulcre J, Dyrba M, Sabri O, Teipel SJ, et al. In vivo staging of regional amyloid deposition. Neurology. 2017 Nov 14;89(20):2031–8. 10.1212/WNL.0000000000004643. PubMed PMID: 29046362.

35. Collij LE, Heeman F, Salvadó G, Ingala S, Altomare D, de Wilde A, et al. Multitracer model for staging cortical amyloid deposition using PET imaging. Neurology. 2020 Sep 15;95(11):e1538–53. 10.1212/WNL.0000000000010256. PubMed PMID: 32675080.

36. Braak H, Braak E. Neuropathological stageing of Alzheimer-related changes. Acta Neuropathol (Berl). 1991;82(4):239–59. 10.1007/BF00308809. PubMed PMID: 1759558.

37. Therriault J, Pascoal TA, Lussier FZ, Tissot C, Chamoun M, Bezgin G, et al. Biomarker modeling of Alzheimer’s disease using PET-based Braak staging. Nat Aging. 2022 Jun;2(6):526–35. 10.1038/s43587-022-00204-0. PubMed PMID: 37118445

38. Pawlik D, Leuzy A, Strandberg O, Smith R. Compensating for choroid plexus based off-target signal in the hippocampus using 18F-flortaucipir PET. NeuroImage. 2020 Nov 1;221:117193. 10.1016/j.neuroimage.2020.117193. PubMed PMID: 32711062.

