## Supplementary Materials for "Biomarker-Informed Interpretation of Dyadic Cognitive Function Index Scores in Cognitively Unimpaired Older Adults"

**Supplementary Methods:**

*Amyloid regional extent categories*

Amyloid burden was quantified using global SUVR [Clark2012; Joshi2015]. For categorical analyses, we constructed A4/LEARN-adapted amyloid regional extent categories based on available regional florbetapir SUVR measures. Because the A4/LEARN quantitative florbetapir dataset provides a limited set of regional SUVR measures, we could not directly implement previously validated amyloid PET staging systems. We therefore constructed A4/LEARN-adapted amyloid regional extent categories, informed by prior PET-based amyloid categorization studies [Mattsson2019;Grothe2017;Collij2020] and constrained by available regional measures. Regional abnormality was defined as SUVR exceeding the 95th percentile of the corresponding regional distribution in the LEARN low-amyloid reference population, consistent with the use of LEARN to preserve the lower-amyloid reference range in A4/LEARN baseline analyses [Sperling2020]. Amyloid regional extent category 0 indicated no abnormal region; amyloid regional extent category 1 indicated abnormality restricted to a predefined early cortical ROI set, consisting of the precuneus, posterior cingulate, and/or medial orbitofrontal cortex; amyloid regional extent category 2 indicated parietal and/or anterior cingulate involvement without temporal involvement or diffuse spread; and amyloid regional extent category 3 indicated broader cortical involvement, defined as temporal involvement and/or diffuse spread across the available regions. Amyloid regional extent category 3 was defined to capture broader cortical involvement within the available A4/LEARN ROI set, not to imply that temporal amyloid deposition universally represents a late event in all amyloid spreading models. These categories should be interpreted as a dataset-specific approximation of regional amyloid extent rather than as a fully validated amyloid regional extent categorization system. Regional cutoffs are provided in Table S1.

Categories were assigned using a mutually exclusive rule-based algorithm designed to approximate regional amyloid extent within the available A4/LEARN ROI set. Participants were first classified as amyloid regional extent category 3 if temporal involvement or diffuse spread across the available cortical ROIs was present. Among the remaining participants, category 2 was assigned if parietal and/or anterior cingulate involvement was present. Category 1 was assigned when abnormality was restricted to the predefined early cortical ROI set, and category 0 indicated no abnormal ROI. The number of participants in each amyloid regional extent category and their demographic and biomarker characteristics are reported in Table S2.

*Tau PET categories and composites*

For the continuous tau PET analysis, we used an early-to-middle temporal tau composite, computed as the mean of standardized (z-scored) bilateral entorhinal and inferior temporal cortex SUVR values. This composite was selected to capture a continuous tau signal spanning early and intermediate tau-vulnerable temporal regions [Schöll2016; Johnson2016; Sanchez2021].

For categorical analyses, we created a simplified PET-based Braak-informed tau PET-based stage using seven bilateral cortical regions [Schöll2016;Braak1991;Therriault2022]. Regional tau abnormality was defined as SUVR exceeding the 95th percentile of the amyloid-negative reference group, defined as florbetapir SUVR < 1.10 or SUVR 1.10–1.15 with a negative visual read. Tau PET-based stage 0 indicated no abnormal region; tau PET-based stage 1 indicated medial temporal involvement only (entorhinal and/or parahippocampal cortex); tau PET-based stage 2 indicated temporal association cortex involvement, defined by inferior temporal and/or fusiform abnormality without association-cortex involvement; and tau PET-based stage 3 indicated neocortical spread (precuneus, posterior cingulate, and/or inferior parietal cortex). This simplified scheme was informed by neuropathological Braak staging and prior tau PET studies, but was adapted to the available A4/LEARN regional tau PET measures. The hippocampus was not included because flortaucipir signal in this region can be affected by adjacent choroid plexus off-target binding [Pawlik2020]. The tau PET composite was standardized within the tau PET available subset (N = 438).

*Plasma p-tau217 assay, transformation, and LLOQ*

Plasma p-tau217, measured at baseline, was standardized (z-scored) within all A4/LEARN participants with available blood data (N = 1,066) to allow comparability with other assays [Palmqvist2020].

***Direct ΔCFI model***

To quantify the degree of individual-level reporter discordance associated with each biomarker, a direct discrepancy score (ΔCFI = CFI-PT − CFI-SP) was computed and regressed on each biomarker separately (amyloid regional extent category, tau PET-based stage, and z-scored plasma p-tau217). Three separate models were estimated, each with a distinct pathology term:

$$\Delta\text{CFI}=\beta_{0}+\beta_{1}Amyloid regional extent category+\beta_{3}\text{Covariates}+\varepsilon$$

$$\Delta\text{CFI}=\beta_{0}+\beta_{1}Tau PET-based stage+\beta_{2}\text{Amyloid }\text{regional extent category}\text{ + }\beta_{3}\text{Covariates}+\varepsilon$$

$$\Delta\text{CFI}=\beta_{0}+\beta_{1}z-pTau217+\beta_{2}\text{Amyloid }\text{regional extent category}\text{ + }\beta_{3}\text{Covariates}+\varepsilon$$

Amyloid regional extent category and tau PET-based stage were modeled as categorical variables (factor, amyloid regional extent category 0 / tau PET-based stage 0 as reference), and plasma p-tau217 was entered as a standardized continuous variable (z-score). Amyloid regional extent category was included as a covariate in the tau PET-based stage and p-tau217 models to isolate the pathology-specific contribution to reporter discordance. Full results are shown in Table S6.

***Continuous amyloid model***

To assess whether the categorical amyloid findings depended on the A4/LEARN-adapted regional extent classification, a complementary analysis was conducted using global florbetapir SUVR as an unstandardized continuous predictor in the long-format interaction model. The key estimand was the global amyloid SUVR × reporter interaction, testing whether the association between continuous amyloid burden and CFI differed between CFI-PT and CFI-SP. Results are shown in Table S5.

*Nested model comparison*

In the tau model, three nested models were estimated. The base model (M1) included amyloid regional extent category, reporter, and their interaction (amyloid regional extent category × reporter), along with covariates. A second model (M2) additionally included the early-to-middle temporal tau composite as a main effect, allowing tau PET burden to contribute to overall complaint burden independently of reporter. A third model (M3) further added the tau × reporter interaction term, testing whether tau shows reporter-specific effects beyond a general burden signal. Nested model comparisons using F tests compared M1 versus M2 (incremental contribution of tau to overall burden) and M2 versus M3 (reporter-specific tau effect).

To examine whether plasma p-tau217 shows a reporter-specific signal within the A4-derived blood biomarker subset, a long-format interaction model was estimated with z-scored plasma p-tau217 as the biomarker term. The interaction term β₃ tests whether plasma p-tau217 differentially predicts CFI-PT versus CFI-SP.

To compare the explanatory value of tau PET and plasma p-tau217 for cross-sectional CFI burden and reporter-specific terms, both biomarkers were evaluated in the matched subset of participants with both measures available. All models included amyloid regional extent category as a covariate alongside age, sex, education, *APOE* ε4 allele count (0, 1, or 2), and PACC. Four models were estimated: a reference amyloid-only model (amyloid regional extent category × reporter + covariates), a tau PET model (amyloid regional extent category × reporter + reporter × early-to-middle temporal tau composite + covariates), a plasma p-tau217 model (amyloid regional extent category × reporter + reporter × z_pTau217 + covariates), and a simultaneous model including both biomarkers with their respective reporter interactions (amyloid regional extent category × reporter + reporter × early-to-middle temporal tau composite + reporter × z_pTau217 + covariates). Nested model comparisons using F tests assessed the incremental contribution of each biomarker beyond the other; the principal model-fit results are summarized in Table 5.

Model fit was compared using AIC and adjusted R². Nested model comparisons using F tests assessed: (i) the incremental contribution of plasma p-tau217 beyond tau PET alone; and (ii) the incremental contribution of tau PET beyond plasma p-tau217 alone.

*Separate-reporter models*

To characterize the direction of any reporter divergence identified in the interaction models, separate ordinary least squares (OLS) regression models were estimated for CFI-PT and CFI-SP as dependent variables, with amyloid and tau entered as predictors in turn, alongside all covariates. These separate-reporter models allow direct inspection of the sign and magnitude of biomarker associations within each reporter.

*Sensitivity analyses*

To assess potential selection associated with biomarker availability, descriptive characteristics were compared across the full baseline sample, the tau PET subsample, the plasma p-tau217 subsample, and the matched subset with both biomarkers available. In the matched subset, nested models were compared to evaluate the incremental contributions of tau PET and plasma p-tau217 to cross-sectional CFI model fit.

*Detailed statistical procedures*

All analyses used listwise deletion. At screening, CFI-PT and CFI-SP were available for 1,685 and 1,683 participants, respectively. Complete-case sample sizes varied slightly across models because of missing covariates. The primary long-format model included 1,682 participants (3,364 observations), whereas the separate-reporter models included 1,683 participants for CFI-PT and 1,681 participants for CFI-SP. When amyloid burden was modeled categorically, amyloid regional extent category was entered as a factor with Amyloid regional extent category 0 as the reference; when modeled continuously, composite amyloid SUVR was entered unstandardized. For categorical amyloid regional extent and tau PET-based stage models, Wald tests were used for the overall pathology × reporter interaction. Category- or stage-specific contrasts were interpreted as exploratory unless supported by the corresponding global interaction test. Findings outside the primary interaction framework were interpreted as supportive rather than definitive.

**Supplementary Results**

The following supplementary results provide supporting analyses for the primary findings reported in the main text.

*Separate-reporter models and direct ΔCFI - amyloid:*

Separate-reporter models supported the direction of this pattern (Table S4): participant report was higher from the restricted early cortical amyloid pattern onward, whereas study partner report was clearly higher only in the temporal/diffuse pattern. These separate-reporter models were used descriptively; formal inference about reporter-specific associations was based on the long-format interaction model. The direct discrepancy analysis showed that only amyloid regional extent category 1 was associated with greater ΔCFI (β = 0.728 on the 0–15 reporter-specific CFI scale, p = 0.004), indicating a statistically detectable but modest participant-leading difference. Adjusted mean CFI scores from the long-format model are shown in Figure 1A. The adjusted within-category PT−SP contrast was largest in the restricted early cortical amyloid pattern, intermediate in the parietal/anterior cingulate pattern, and attenuated in the temporal/diffuse pattern, visually supporting the interaction-based finding that participant-leading imbalance was most apparent in the restricted early cortical amyloid pattern. Neither tau PET-based stage nor plasma p-tau217 showed significant associations with ΔCFI. Full results are shown in Table S6.

*Continuous amyloid model:*

In the complementary continuous amyloid model, global florbetapir SUVR showed no clear reporter interaction pattern (global amyloid SUVR × study partner report: β = -0.391, SE = 0.269, p = 0.146; Table S5). This analysis was used to assess whether the categorical amyloid findings depended on the A4/LEARN-adapted regional extent classification. The absence of a continuous global SUVR × reporter interaction suggests that the participant-leading pattern was more apparent when amyloid was represented as regional extent rather than as global burden alone.

*Tau PET stage-based separate-reporter models:*

Descriptive stage-based separate-reporter models did not show a simple monotonic increase across tau PET-based stages (Table S7). Stage 2 was associated with lower CFI-PT relative to stage 0, whereas stage 3 showed a positive association with CFI-PT and a weaker, non-significant positive association with CFI-SP. Because these stage-based findings were descriptive and the formal continuous tau × reporter interaction was not significant, they should not be interpreted as evidence of reporter-specific tau-related divergence. Adjusted mean CFI scores are shown in Figure 1B, illustrating broadly parallel reporter differences across tau PET-based stages. Descriptive scatterplots for continuous amyloid burden and early-to-middle temporal tau composite burden are provided in Figure S2. Thus, the primary tau analysis supports an association with overall CFI burden, but provides no evidence that tau-CFI associations differed between reporters.

*Sensitivity Analyses:*

Sample availability analyses indicated that the tau PET subsample represented a selected subset of the full A4/LEARN sample, with higher amyloid positivity and greater A4 representation (Table S3). The plasma p-tau217 subset was entirely A4-derived and did not include LEARN participants. Results from the tau PET and plasma p-tau217 analyses should therefore be interpreted in light of these biomarker-specific sample restrictions. Model comparisons within the matched subset are reported in Table 5.

**Table S1. Regional florbetapir SUVR cutoffs and rule-based biomarker category definitions**

Table S1A. Regional florbetapir SUVR cutoffs used for amyloid regional extent categories

| **Region group** | **Region** | **Variable label** | **Cutoff source** | **Cutoff (SUVR)** | **Abnormality rule** | **Role in amyloid regional extent category assignment** |
| --- | --- | --- | --- | --- | --- | --- |
| Restricted early cortical | Precuneus | *lprecuneus_gm* | 95th percentile in LEARN reference population | 1.23 | SUVR > cutoff | Predefined early cortical ROIs used to define restricted early regional extent |
|  | Posterior cingulate | *llposterior_cingulate_2* | 95th percentile in LEARN reference population | 1.21 | SUVR > cutoff | Predefined early cortical ROIs used to define restricted early regional extent |
|  | Medial orbitofrontal | *xlaal_frontal_med_orb* | 95th percentile in LEARN reference population | 1.02 | SUVR > cutoff | Predefined early cortical ROIs used to define restricted early regional extent |
| Intermediate/regional extension | Parietal | *lnew_parietal* | 95th percentile in LEARN reference population | 1.14 | SUVR > cutoff | Parietal/anterior cingulate ROI used to define intermediate regional extent |
|  | Anterior cingulate | *lanterior_cingulate_2* | 95th percentile in LEARN reference population | 1.19 | SUVR > cutoff | Parietal/anterior cingulate ROI used to define intermediate regional extent |
| Late/broad cortical extent | Temporal | *new_temporal_2* | 95th percentile in LEARN reference population | 1.18 | SUVR > cutoff | Temporal ROI used to define broad cortical regional extent |

Table S1B. Amyloid regional extent category algorithm

| **Amyloid regional extent category** | **Operational definition for manuscript** | **Interpretation / note** |
| --- | --- | --- |
| Category 0 | No abnormal ROI among the six available amyloid ROIs. | Reference category. |
| Category 1 | One or more predefined early cortical ROIs are abnormal, with no parietal/anterior cingulate and no temporal involvement. | Early restricted regional extent. |
| Category 2 | Parietal and/or anterior cingulate involvement without temporal involvement and without diffuse spread across ≥5 ROIs. | This category does not require early ROI positivity in the code; avoid wording that implies a strictly hierarchical temporal sequence. |
| Category 3 | Temporal involvement and/or diffuse spread across five or more of the six available amyloid ROIs. | Broad cortical regional extent; not a validated late amyloid regional extent category. |

Table S1C. Tau PET continuous measure and tau PET-based stage algorithm

| **Measure / tau PET-based stage** | **Operational definition for manuscript** | **Interpretation / note** |
| --- | --- | --- |
| Early-to-middle temporal tau composite | Mean of z-scored bilateral entorhinal and inferior temporal SUVR values in the tau PET subset. | Primary continuous tau PET measure. |
| Tau PET-based stage 0 | No abnormal tau ROI among the seven available tau PET ROIs. | Reference tau PET-based stage. |
| Tau PET-based stage 1 | Medial temporal involvement only: entorhinal and/or parahippocampal cortex. | Braak-informed early tau PET-based stage. |
| Tau PET-based stage 2 | Temporal association cortex involvement: inferior temporal and/or fusiform cortex, with no association-cortex involvement. | The code does not require medial temporal positivity for tau PET-based stage 2. |
| Tau PET-based stage 3 | Association/neocortical involvement: precuneus, posterior cingulate, and/or inferior parietal cortex. | Highest tau PET-based stage in this simplified A4/LEARN-adapted scheme. |

Amyloid regional abnormality was defined as SUVR exceeding the 95th percentile of the corresponding regional florbetapir SUVR distribution in the LEARN reference population. Amyloid regional extent categories were assigned using mutually exclusive rule-based criteria adapted to the available A4/LEARN regional SUVR measures. These categories should be interpreted as dataset-specific approximations of regional amyloid extent, not as a fully validated temporal sequence of amyloid spread. Tau regional abnormality was defined as SUVR exceeding the 95th percentile of the amyloid-negative reference group. The hippocampus was not included in the tau PET-based stage algorithm because flortaucipir signal in this region can be affected by adjacent choroid plexus off-target binding.

**Abbreviations**: LEARN, Longitudinal Evaluation of Amyloid Risk and Neurodegeneration; PET, positron emission tomography; ROI, region of interest; SUVR, standardized uptake value ratio.

**Table S2. Amyloid regional extent category-specific characteristics**

| **Variable** | **Category 0** | **Category 1** | **Category 2** | **Category 3** | **Overall** |
| --- | --- | --- | --- | --- | --- |
| N | 473 | 87 | 215 | 911 | 1686 |
| A4, N (%) | 41 (8.7%) | 44 (50.6%) | 171 (79.5%) | 892 (97.9%) | 1148 (68.1%) |
| LEARN, N (%) | 432 (91.3%) | 43 (49.4%) | 44 (20.5%) | 19 (2.1%) | 538 (31.9%) |
| Global amyloid SUVR (mean +/- SD) | 0.98 ± 0.06 | 1.1 ± 0.05 | 1.15 ± 0.07 | 1.38 ± 0.17 | 1.22 ± 0.22 |
| *APOE* ε4 carriers (≥1 allele), N (%) | 99 (20.9%) | 25 (28.7%) | 105 (48.8%) | 571 (62.7%) | 800 (47.4%) |
| *APOE* ε4 homozygous, N (%) | 0 (0%) | 0 (0%) | 13 (6%) | 83 (9.1%) | 96 (5.7%) |
| Age (years, mean +/- SD) | 70.4 ± 4.3 | 70.4 ± 4.2 | 70.5 ± 4.2 | 72.3 ± 4.8 | 71.5 ± 4.7 |
| Sex (Male), N (%) | 187 (39.5%) | 19 (21.8%) | 100 (46.5%) | 368 (40.4%) | 674 (40%) |
| Education (years, mean +/- SD) | 16.7 ± 2.6 | 16.4 ± 2.7 | 16.7 ± 3.1 | 16.6 ± 2.8 | 16.6 ± 2.8 |
| PACC raw score (mean +/- SD) | 0.9 ± 2.3 | 0.5 ± 2.4 | 0.5 ± 2.6 | -0.1 ± 2.7 | 0.3 ± 2.6 |
| CFI-PT (mean +/- SD) | 1.7 ± 1.8 | 2.4 ± 2 | 2.4 ± 2.6 | 2.4 ± 2.1 | 2.2 ± 2.1 |
| CFI-SP (mean +/- SD) | 1.1 ± 1.6 | 0.9 ± 1.5 | 1.4 ± 1.9 | 1.6 ± 2 | 1.4 ± 1.9 |
| Delta CFI = CFI-PT - CFI-SP (mean +/- SD) | 0.6 ± 1.9 | 1.5 ± 1.9 | 1 ± 2.4 | 0.8 ± 2.3 | 0.8 ± 2.2 |

Baseline demographic, cognitive, biomarker, and CFI characteristics are shown by A4/LEARN-adapted amyloid regional extent category. Values are mean ± SD for continuous variables and N (%) for categorical variables. Amyloid regional extent category 0 indicates no abnormal ROI; category 1 indicates abnormality restricted to restricted early cortical regions; category 2 indicates parietal and/or anterior cingulate involvement without temporal involvement or diffuse spread; and category 3 indicates temporal involvement and/or diffuse spread across available cortical ROIs. Positive ΔCFI indicates a participant-leading pattern.

**Abbreviations**: A4, Anti-Amyloid Treatment in Asymptomatic Alzheimer’s Disease; APOE, apolipoprotein E; CFI, Cognitive Function Index; CFI-PT, CFI participant report; CFI-SP, CFI study partner report; ΔCFI, discordance score; LEARN, Longitudinal Evaluation of Amyloid Risk and Neurodegeneration; PACC, Preclinical Alzheimer Cognitive Composite; ROI, region of interest; SD, standard deviation; SUVR, standardized uptake value ratio.

**Table S3. Demographic and biomarker characteristics across analytic subsamples**

| **Variable** | **Full sample (N=1686)** | **Tau PET subsample (N=438)** | **p-tau217 subsample (N=1066)** | **Matched subset (N=352)** |
| --- | --- | --- | --- | --- |
| N | 1686 | 438 | 1066 | 352 |
| A4, N (%) | 1148 (68.1%) | 383 (87.4%) | 1066 (100%) | 352 (100%) |
| LEARN, N (%) | 538 (31.9%) | 55 (12.6%) | 0 (0%) | 0 (0%) |
| Age (years, mean +/- SD) | 71.5 ± 4.7 | 71.8 ± 4.8 | 71.9 ± 4.8 | 72.1 ± 4.8 |
| Sex (Male), N (%) | 674 (40%) | 184 (42%) | 441 (41.4%) | 150 (42.6%) |
| Education (years, mean +/- SD) | 16.6 ± 2.8 | 16.2 ± 2.9 | 16.5 ± 2.8 | 16.2 ± 2.8 |
| *APOE* ε4 carriers (≥1 allele), N (%) | 800 (47.4%) | 232 (53%) | 633 (59.4%) | 199 (56.5%) |
| *APOE* ε4 homozygous, N (%) | 96 (5.7%) | 27 (6.2%) | 87 (8.2%) | 25 (7.1%) |
| Amyloid positive, N (%) | 1091 (64.7%) | 356 (81.3%) | 1014 (95.1%) | 327 (92.9%) |
| Global amyloid SUVR (mean +/- SD) | 1.22 ± 0.22 | 1.27 ± 0.19 | 1.33 ± 0.18 | 1.31 ± 0.17 |
| PACC raw score (mean +/- SD) | 0.3 ± 2.6 | -0.1 ± 2.8 | 0 ± 2.7 | -0.2 ± 2.8 |
| CFI-PT (mean +/- SD) | 2.2 ± 2.1 | 2.4 ± 2.2 | 2.3 ± 2.2 | 2.4 ± 2.2 |
| CFI-SP (mean +/- SD) | 1.4 ± 1.9 | 1.5 ± 1.9 | 1.5 ± 2 | 1.5 ± 2 |
| Delta CFI (PT - SP, mean +/- SD) | 0.8 ± 2.2 | 0.8 ± 2.4 | 0.9 ± 2.3 | 0.8 ± 2.4 |
| Plasma p-tau217 among available cases (mean +/- SD) | 0.28 ± 0.16 | 0.27 ± 0.15 | 0.28 ± 0.16 | 0.27 ± 0.15 |

Baseline characteristics are shown for the full A4/LEARN sample and three biomarker-defined analytic subsamples. The tau PET subsample included participants with flortaucipir PET available. The plasma p-tau217 subsample included A4-derived participants with available blood biomarker data. The matched subset included participants with both tau PET and plasma p-tau217 available. Values are mean ± SD for continuous variables and N (%) for categorical variables. Positive ΔCFI indicates a participant-leading pattern.

**Abbreviations**: A4, Anti-Amyloid Treatment in Asymptomatic Alzheimer’s Disease; APOE, apolipoprotein E; CFI, Cognitive Function Index; CFI-PT, CFI participant report; CFI-SP, CFI study partner report; ΔCFI, CFI-PT minus CFI-SP; LEARN, Longitudinal Evaluation of Amyloid Risk and Neurodegeneration; PACC, Preclinical Alzheimer Cognitive Composite; PET, positron emission tomography; p-tau217, phosphorylated tau 217; SUVR, standardized uptake value ratio.

**Table S4. Separate-reporter OLS models: CFI-PT and CFI-SP as dependent variables**

| **label** | **CFI-PT beta** | **CFI-PT SE** | **CFI-PT p-value** | **CFI-SP beta** | **CFI-SP SE** | **CFI-SP p-value** |
| --- | --- | --- | --- | --- | --- | --- |
| Amyloid regional extent category 1 (vs category 0) | 0.608 | 0.242 | 0.012 | -0.095 | 0.219 | 0.663 |
| Amyloid regional extent category 2 (vs category 0) | 0.561 | 0.173 | 0.001 | 0.216 | 0.156 | 0.166 |
| Amyloid regional extent category 3 (vs category 0) | 0.420 | 0.130 | 0.001 | 0.394 | 0.117 | <0.001*** |

Separate-reporter models were refit with explicit treatment coding for amyloid regional extent category, using category 0 as the reference. CFI-PT N = 1,683; CFI-SP N = 1,681. The N differs slightly from the number of available CFI observations because separate-reporter models used listwise deletion for all covariates. These models were used to characterize the direction of associations within each reporter; formal inference about reporter-specific effects was based on the long-format interaction model in Table 2. All models were adjusted for age, sex, years of education, APOE ε4 allele count, and baseline cognition (PACC).

**Abbreviations**: β, regression coefficient; CFI-PT, Cognitive Function Index participant report; CFI-SP, Cognitive Function Index study partner report; OLS, ordinary least squares; * p < 0.05; ** p < 0.01; *** p < 0.001.

**Table S5. Continuous amyloid model: global florbetapir SUVR × reporter interaction**

| **label** | **beta** | **SE** | **p-value** | **sig** |
| --- | --- | --- | --- | --- |
| Intercept | -0.038 | 0.800 | 0.962 |  |
| Global amyloid SUVR (continuous) | 1.319 | 0.259 | <0.001 | *** |
| Reporter: CFI-SP (ref = CFI-PT) | -0.335 | 0.321 | 0.297 |  |
| Age | 0.010 | 0.009 | 0.273 |  |
| Female sex (1 = female; reference = male) | -0.209 | 0.090 | 0.020 | * |
| Education (years) | 0.002 | 0.016 | 0.885 |  |
| *APOE* ε4 allele count (0, 1, or 2) | -0.004 | 0.070 | 0.955 |  |
| PACC raw score | -0.097 | 0.017 | <0.001 | *** |
| Global amyloid SUVR × CFI-SP | -0.391 | 0.269 | 0.146 |  |

Complementary analysis to the primary categorical amyloid model. Global florbetapir SUVR was entered as an unstandardized continuous predictor in a long-format interaction model with two observations per participant. The key estimand is the global amyloid SUVR × reporter interaction, which tests whether the association between continuous amyloid burden and CFI differs between CFI-PT and CFI-SP. The model was refit with explicit treatment coding, with CFI-PT as the reporter reference, so the reporter and interaction terms are interpretable as CFI-SP minus CFI-PT effects. All models were adjusted for age, sex, education, APOE ε4 allele count, and PACC. Standard errors were clustered by participant.

**Abbreviations**: APOE, apolipoprotein E; CFI, Cognitive Function Index; CFI-PT, CFI participant report; CFI-SP, CFI study partner report; PACC, Preclinical Alzheimer Cognitive Composite; SE, standard error; SUVR, standardized uptake value ratio.

**Table S6. Full results of direct CFI discordance models**

| **Variable** | **Amyloid regional extent category: B (SE, p-value)** | **Tau PET-based stage: B (SE, p-value)** | **p-tau217: B (SE, p-value)** |
| --- | --- | --- | --- |
| Intercept | 0.458 (SE=0.958, p=0.633) | 0.503 (SE=2.016, p=0.803) | 0.081 (SE=1.279, p=0.949) |
| Amyloid regional extent category 1 (ref = category 0) | 0.728 (SE=0.255, p=0.004**) | 1.650 (SE=0.611, p=0.007**) | 0.791 (SE=0.522, p=0.130) |
| Amyloid regional extent category 2 (ref = category 0) | 0.344 (SE=0.181, p=0.058.) | 0.294 (SE=0.436, p=0.500) | 0.077 (SE=0.421, p=0.856) |
| Amyloid regional extent category 3 (ref = category 0) | 0.025 (SE=0.137, p=0.855) | -0.159 (SE=0.367, p=0.665) | -0.227 (SE=0.400, p=0.570) |
| Age | 0.013 (SE=0.013, p=0.290) | 0.016 (SE=0.026, p=0.546) | 0.016 (SE=0.016, p=0.313) |
| Female sex (1 = female; reference = male) | 0.490 (SE=0.113, p=<0.001***) | 0.315 (SE=0.246, p=0.201) | 0.379 (SE=0.149, p=0.011*) |
| Education | -0.064 (SE=0.020, p=0.001**) | -0.076 (SE=0.042, p=0.068.) | -0.039 (SE=0.026, p=0.128) |
| *APOE* ε4 allele count (0, 1, or 2) | 0.164 (SE=0.097, p=0.090.) | 0.161 (SE=0.206, p=0.437) | 0.227 (SE=0.118, p=0.055.) |
| PACC raw score | -0.014 (SE=0.023, p=0.538) | -0.028 (SE=0.046, p=0.538) | -0.034 (SE=0.029, p=0.237) |
| Tau PET-based stage 1 (ref = stage 0) |  | -0.272 (SE=0.497, p=0.584) |  |
| Tau PET-based stage 2 (ref = stage 0) |  | 0.233 (SE=0.373, p=0.532) |  |
| Tau PET-based stage 3 (ref = stage 0) |  | 0.478 (SE=0.279, p=0.088.) |  |
| Plasma p-tau217 (z-score) |  |  | 0.019 (SE=0.075, p=0.800) |

Three separate ordinary least squares regression models were estimated with ΔCFI = CFI-PT − CFI-SP as the outcome. Positive ΔCFI indicates a participant-leading pattern. Model 1 tested amyloid regional extent category as the primary predictor. Model 2 tested tau PET-based stage as the primary predictor, with amyloid regional extent category included as a covariate. Model 3 tested z-scored plasma p-tau217 as the primary predictor, with amyloid regional extent category included as a covariate. All categorical predictors were refit with explicit treatment coding, using category or stage 0 as the reference.

**Abbreviations**: APOE, apolipoprotein E; CFI, Cognitive Function Index; CFI-PT, CFI participant report; CFI-SP, CFI study partner report; ΔCFI, CFI-PT minus CFI-SP; PACC, Preclinical Alzheimer Cognitive Composite; PET, positron emission tomography; p-tau217, phosphorylated tau 217; SE, standard error. *** p < 0.001; ** p < 0.01; * p < 0.05; . p < 0.10.

**Table S7. Separate-reporter OLS models: CFI-PT and CFI-SP as dependent variables, tau PET-based stage predictor**

| **label** | **CFI-PT beta** | **CFI-PT SE** | **CFI-PT p-value** | **CFI-SP beta** | **CFI-SP SE** | **CFI-SP p-value** |
| --- | --- | --- | --- | --- | --- | --- |
| Tau PET-based stage 1 (vs stage 0) | -0.276 | 0.179 | 0.124 | -0.170 | 0.158 | 0.281 |
| Tau PET-based stage 2 (vs stage 0) | -0.741 | 0.325 | 0.023 | -0.366 | 0.286 | 0.201 |
| Tau PET-based stage 3 (vs stage 0) | 0.505 | 0.251 | 0.045 | 0.377 | 0.221 | 0.089 |

Separate ordinary least squares regression models estimated for CFI-PT and CFI-SP as dependent variables, with tau PET-based stage as the primary predictor, in the tau PET subsample (N = 438). Tau PET-based stage 0 was the reference category. These models were used descriptively to characterize the direction of associations within each reporter; formal inference about reporter-specific divergence was based on the tau × reporter interaction term in the long-format model (Table 3). All models were adjusted for amyloid regional extent category, age, sex, years of education, APOE ε4 allele count, and baseline cognition (PACC).

**Abbreviations**: CFI-PT, Cognitive Function Index participant report; CFI-SP, Cognitive Function Index study partner report; OLS, ordinary least squares; PACC, Preclinical Alzheimer Cognitive Composite; PET, positron emission tomography; SE, standard error

**Table S8. Nested model comparisons**

| **comparison** | **F** | **df** | **p-value** | **Delta R2** |
| --- | --- | --- | --- | --- |
| Amyloid-only vs + early-to-middle temporal tau composite | 17.49 | 1 | <0.001 | 0.018 |
| + tau composite main effect vs + tau × reporter | 0.05 | 1 | 0.817 | 0.000 |

Nested F tests comparing long-format models of increasing complexity in the tau PET subsample (N = 438 participants; 876 long-format observations). The first comparison tested whether the early-to-middle temporal tau composite improved model fit beyond the baseline amyloid regional extent category × reporter model. The second comparison tested whether adding the tau × reporter interaction improved fit beyond the tau main-effect model. All models were adjusted for age, sex, years of education, APOE ε4 allele count, and baseline cognition (PACC). Full model coefficients are shown in Table 3.

**Abbreviations**: PACC, Preclinical Alzheimer Cognitive Composite; PET, positron emission tomography; R², coefficient of determination.

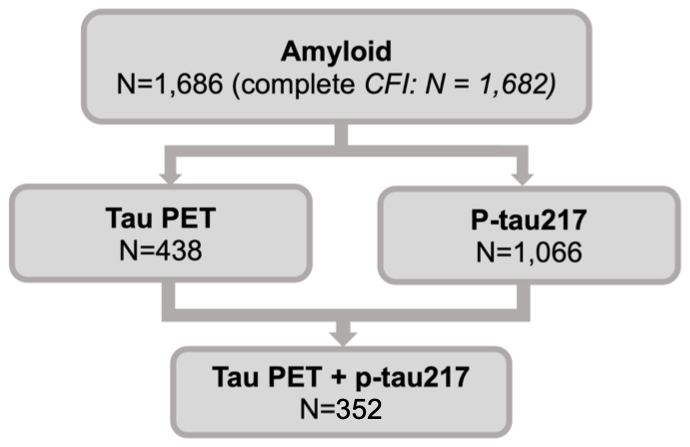
**Figure S1:** **Analytic subsample availability across biomarker configurations**

The figure summarizes biomarker availability in the A4/LEARN screening sample. The full sample included 1,686 cognitively unimpaired participants with amyloid PET available, of whom 1,682 had complete dyadic CFI data for the primary long-format analyses. Tau PET was available for 438 participants, plasma p-tau217 for 1,066 participants, and both tau PET and plasma p-tau217 for 352 participants.

**Abbreviations**: A4, Anti-Amyloid Treatment in Asymptomatic Alzheimer’s Disease; CFI, Cognitive Function Index; LEARN, Longitudinal Evaluation of Amyloid Risk and Neurodegeneration; PET, positron emission tomography; p-tau217, phosphorylated tau 217.

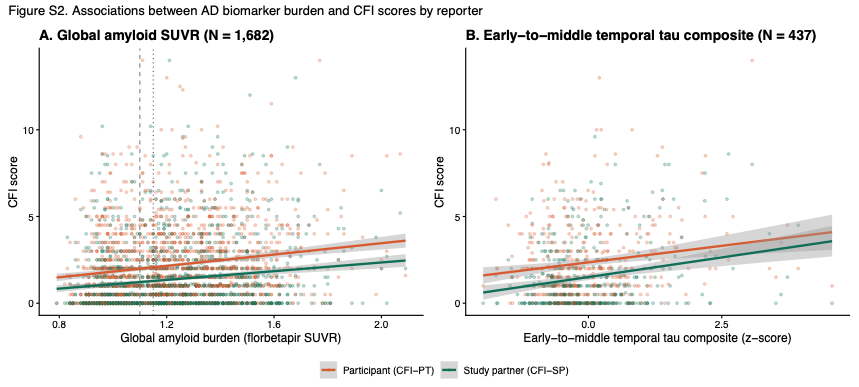
**Figure S2. Associations between AD biomarker burden and CFI scores by reporter**

Scatterplots show participant-reported and study partner-reported CFI scores as a function of global amyloid burden and early-to-middle temporal tau composite burden. Regression lines and 95% confidence intervals are shown separately by reporter. Panel A uses the full dyadic CFI sample with amyloid data. Panel B is restricted to participants with tau PET available. These plots are descriptive and complement the primary long-format interaction models.

**Abbreviations**: AD, Alzheimer’s disease; CFI, Cognitive Function Index; CFI-PT, CFI participant report; CFI-SP, CFI study partner report; CI, confidence interval; PET, positron emission tomography; SUVR, standardized uptake value ratio.
